# Effect of Boysenberry apple powder blend (BerriQi^®^) on reducing symptom severity in children with Upper respiratory tract infection

**DOI:** 10.64898/2026.07.26.26358747

**Authors:** Aahana Shrestha, Emma Graham, Starin Mckeen

## Abstract

**Background/Objective:** Seasonal upper respiratory tract infections (URTIs) are common in children and contribute to absenteeism and reduced quality of life. Over-the-counter treatments show limited efficacy and may cause adverse effects, thus warranting the need for natural alternatives. This study evaluated the efficacy of a whole-fruit supplement derived from boysenberry and apple (BerriQi) in reducing symptom severity and duration in school-aged children with URTIs using the Wisconsin Upper Respiratory Symptoms Survey for Kids (WURSS-K).

**Methods:** In this double-blind, randomised, placebo-controlled study, 84 children aged 5–13 years with URTI-associated school absence were assigned to receive BerriQi or placebo (two chewable tablets daily) for 14 days. Outcomes included the severity and duration of global illness, composite symptom and function scores.

**Results:** Both groups showed progressive reductions in global illness severity, symptom, and function score over 14 days. However, the BerriQi group consistently reported lower scores across all outcomes. While global illness severity did not differ significantly between groups, total symptom severity was significantly reduced with BerriQi (P=0.04), and functional scores showed a trend toward improvement (P=0.05). The BerriQi group experienced fewer sick days compared with placebo (8 vs. 11 days) and demonstrated a higher likelihood of functional recovery (HR = 1.79, 95% CI: 1.09– 2.91; P=0.02).

**Conclusions:** These findings suggest that BerriQi may serve as a promising natural paediatric supplement for alleviating respiratory illness symptoms and supporting faster functional recovery, potentially reducing school absenteeism associated with upper respiratory tract infections (URTIs).

## 1 Introduction

The common cold, a viral upper respiratory tract infection (URTI), is the most common acute illness in childhood and a major cause of absenteeism from school and work (Pappas, 2017). Children aged 5-9 years’ experience largest episodes of URTI (Sirota et al., 2025), which contribute substantially to healthcare utilisation, parental work loss and school absenteeism (Schot et al., 2019), particularly during colder months when respiratory infections are more prevalent (Tang et al., 2025). In New Zealand, national education data indicate higher school absenteeism during terms two and three (April– September), coinciding with peak circulation of seasonal respiratory pathogens (Ministry of Education, 2022). These non-influenza viral respiratory tract infections incur a high cost in both direct healthcare expenditures and indirect costs associated with absenteeism from school and work; in the USA, this annual cost exceeds 40 billion dollars (Fendrick et al, 2003), rising to $170 billion when all respiratory diseases are included (Nurmagambetov, 2022).

The treatment for URTIs mainly focuses on symptom relief that involves over-the-counter medications such as antipyretics, decongestants and nonsteroidal anti-inflammatory drugs, yet there is limited evidence that these therapies reduce illness duration. Moreover, several of these medications have not demonstrated clear benefit over placebo in paediatric populations and may be associated with adverse effects (Cotton et al 2011). Dietary supplements, especially vitamin C, have been suggested as a means of preventing the common cold, however, findings across studies have been inconsistent. A randomised controlled study with low and high dose vitamin C over a 5-year period in Japan found no reduction in the duration or severity of the common cold, although the study suggested a reduction in the frequency of common cold episodes (Sasazuki et al., 2006). Similarly, different parts of elderberry have been used in traditional practice for respiratory illness, but its use does not reduce the incidence of colds. A recent systematic review suggested that elderberries may reduce the duration and severity of colds, although the certainty of the evidence was low (Wieland et al., 2021). With respect to safety, the European Medicines Agency has advised that only the flowers and fully ripened berries are suitable for consumption, and that adequate heating of the fruit or juice is recommended prior to use (Gro et al., 2014). To the best of our knowledge, few evidence-based, child-appropriate, non- pharmaceutical interventions offering consistent and reliable symptomatic relief are currently available for paediatric populations. There is therefore an unmet, growing clinical need for safe, natural therapeutic options that can reduce both the severity and duration of URTI symptoms.

Epidemiological evidence supports a positive correlation between high fruit and vegetable consumption and improved pulmonary function, as well as a reduced risk of chronic obstructive pulmonary disease (COPD) and asthma (Kaluza et al., 2018; Park and Park, 2025) These benefits are largely attributed to the high concentrations of vitamins, minerals, and phenolic compounds in fruits and vegetables, which collectively confer antioxidant and anti-inflammatory properties (Emran et al., 2024).

BerriQi^®^ is a whole-fruit formulation derived from New Zealand Boysenberry and apple (naturally rich in anthocyanins, quercetin and procyanidins) that has demonstrated efficacy in preclinical models of allergic airway inflammation, reducing mucus production, immune cell infiltration, and airway tissue damage, thereby improving lung function (Shaw et al., 2020). Despite these mechanistic and *in vivo* data, BerriQi has yet to be evaluated in the context of acute viral URTIs. Given its established preclinical effects on airway inflammation, it is plausible that BerriQi may attenuate the severity and duration of respiratory symptoms in paediatric URTIs and thereby influence functional outcomes such as school attendance. Therefore, the present study was designed to assess whether BerriQi reduces the perceived severity and duration of URTI symptoms in school aged children, using the validated Wisconsin Upper Respiratory Symptoms Survey for Kids (WURSS-K).

## 2 Materials and Methods

### 2.1 Study Designs and Procedures

This was a randomised, double-blind, placebo-controlled parallel trial evaluating the effect of BerriQi®, a boysenberry and apple blend, on post-respiratory-infection recovery among primary-school children in New Zealand. The trial was registered with the Australia New Zealand Clinical Trials Registry (registration number ACTRN: 12624000398505).

#### 2.1.1 Recruitment

Participants were recruited New Zealand-wide through social media advertisements from 1^st^ August 2024 to 5^th^ August 2025. Interested families (caregivers) of potential participants were informed about the study and provided written consent. Families and caregivers were encouraged to include their children in the decision to participate in the study and the children also completed assent forms. Only children for whom both consent and assent forms had been completed, and who subsequently developed a URTI leading to school absence, were randomised to the intervention and enrolled in the study.

#### 2.1.2 Participants

Eligible participants were children aged 5–13 years who reported URTI symptoms, whose caregivers provided informed consent, and who provided informed assent. Exclusion criteria were allergy to berries or fruits and any condition judged by investigators to interfere with participation.

### 2.2 Randomisation and blinding

Participants were randomised in a 1:1 ratio to receive either BerriQi® or placebo using a randomisation sequence generated in SAS software and implemented through REDCap (University of Auckland). Variable block sizes were used to maintain concealment. Randomisation and product allocation were performed by a non-research staff member. Investigators, caregivers, school staff, statisticians, and outcome assessors remained blinded until the database was unlocked on 29 October 2025.

### 2.3 Interventions

The interventions comprised either BerriQi® or placebo chewables. BerriQi is a freeze-dried powder derived from Boysenberries and apples. For placebo, the Boysenberry component was heat-treated to remove heat-sensitive actives, but the apple component remained the same. Each chewable was composed of 500 mg of BerriQi, 417 mg xylitol, and 33 mg magnesium stearate. The placebo chewable had the same composition except the Boysenberry component was heated prior to the freeze-drying process. Participants took two chewable tablets (active or placebo) daily for 14 days. The active treatment delivered 6.4 mg of actives (anthocyanins, quercetin and procyanidins) per day (3.2 mg per tablet), compared with 0.5 mg of actives (anthocyanins, quercetin and procyanidins) per day in the placebo (i.e., 0.25 mg per tablet). The active and placebo chewables were matched for shape, colour, and size.

### 2.4 Outcome Measure

Wisconsin Upper Respiratory Symptoms Survey for Kids (WURSS-K) is a validated and reliable illness-specific quality of life instrument focusing on both illness-specific symptoms and their impact on quality of life (Schmit et al., 2021). It is a shorter and simpler version of the WURSS questionnaires specifically designed for children aged 4-10 years to evaluate the impact of upper respiratory infections on children. It includes 15 items covering global illness severity (item 1), severity of symptoms (items 2-7; i.e., runny nose, stuffy nose, sneezing, sore throat, cough and feeling tired), functional impacts (items 8-14; i.e. thinking, sleeping, breathing, talking, walking/climbing stairs/exercise, going to school, and playing with friends), and evaluation of change over time (item 15) (Supplementary Figure 1). Happy- and sad-face ordinal scale representations were included to help survey completion by children. However, parental help may have been required, especially for children below 8 years of age.

#### 2.4.1 Primary outcome

The primary aim of the study was to investigate whether BerriQi improved the severity of daily global illness severity score (WURSS-K item 1: “How sick do you feel today?”; 1 = not sick, 4 = very bad) over 14 days compared to placebo.

#### 2.4.2 Secondary outcomes

The study measured whether BerriQi improved symptom severity and functional symptoms across 14 days compared to placebo. Symptom sum (items 2-7 i.e., runny nose, stuffy nose, sneezing, sore throat, cough, and feeling tired; range 0–18). Function sum (items 8-14; i.e., thinking, sleeping, breathing, talking, walking/climbing stairs/exercise, going to school, and playing with friends; range 0–21).

Additionally, the study compared total sick days and symptom resolution using both the global illness severity score and the symptom sum score. Time to symptom resolution was calculated based on the following criteria:

- First occurrence of two consecutive days with global illness score = 1 (“not sick”)
- First occurrence of two consecutive days with symptom sum = 6 (corresponding to “not sick” for all the symptoms measured)

### 2.5 Sample size calculations

Based on Barrett et al. (2005) using the WURSS, an effect size of 0.709 was expected. A total of 52 participants (26 per group) provided 80 % power at α = 0.05 (two-sided). Allowing for 20% attrition, the target sample size was 62 participants.

#### 2.5.1 Analysis populations

Intention-to-treat (ITT) included all randomised participants, analysed by assigned group regardless of product receipt or diary completion.

Per-protocol (PP) included participants with ≥ 80 % product-consumption days and ≥ 10 of 14 daily WURSS-K entries.

### 2.6 Statistical Analysis

#### 2.6.1 Primary analysis

Daily global illness scores were analysed using a linear mixed-effects model with fixed effects for treatment group (BerriQi vs placebo), day (1–14), and their interaction, plus a random intercept for participant. Baseline global illness score (day 0) was included as a covariate.

The treatment effect was expressed as the adjusted mean difference in global score with 95% confidence interval (CI). Day-by-day post-hoc contrasts were performed with Tukey-HSD, and p-values were adjusted using the false discovery rate (FDR) method for multiplicity. All analyses followed the intention-to-treat principle with two-sided α = 0.05. Continuous data were summarised as mean ± SEM (or median with confidence interval).

#### 2.6.2 Secondary analyses

Symptom-sum and function-sum scores were modelled identically to the primary outcome.

Days absent from school were compared using negative binomial regression, expressed as incidence-rate ratios (IRR [95% CI]).

Time-to-resolution was analysed by Kaplan–Meier curves, log-rank test, and Cox proportional-hazards model, reported as hazard ratios (HR [95 % CI]).

##### Sensitivity analyses

Per-protocol (PP) analysis repeated the primary model among compliant participants.

##### Cluster handling

Participants from the same family (siblings) were to be treated as clusters by adding a random intercept for family ID when ≥ 2 families had multiple children enrolled.

Analyses were performed in R (version 4.4.1) using the packages dplyr, lme4, lmerTest, emmeans, survival, MASS, and gtsummary.

##### Compliance

Compliance was defined as ≥ 80 % of product-consumption days and ≥ 10/14 survey days. Compliance was summarised per participant and used to classify the PP population.

## 3 Results

### 3.1 Study population

As detailed in Figure 1, a total of 245 children were screened between 1^st^ August 2024 and 5 August 2025. A total of 84 participants were randomised (Treatment, BerriQi, n = 42; Treatment 2, placebo, n = 42). Three children (in BerriQi group) did not receive the allocated product because the product was not delivered before symptom resolution. Eighty-one participants completed at least one daily diary and were included in the outcome analyses. Sixty-four participants (81 %) met the per-protocol criteria (≥ 80 % product use and ≥ 10/14 diary days, Figure 1).

**Figure 1.**
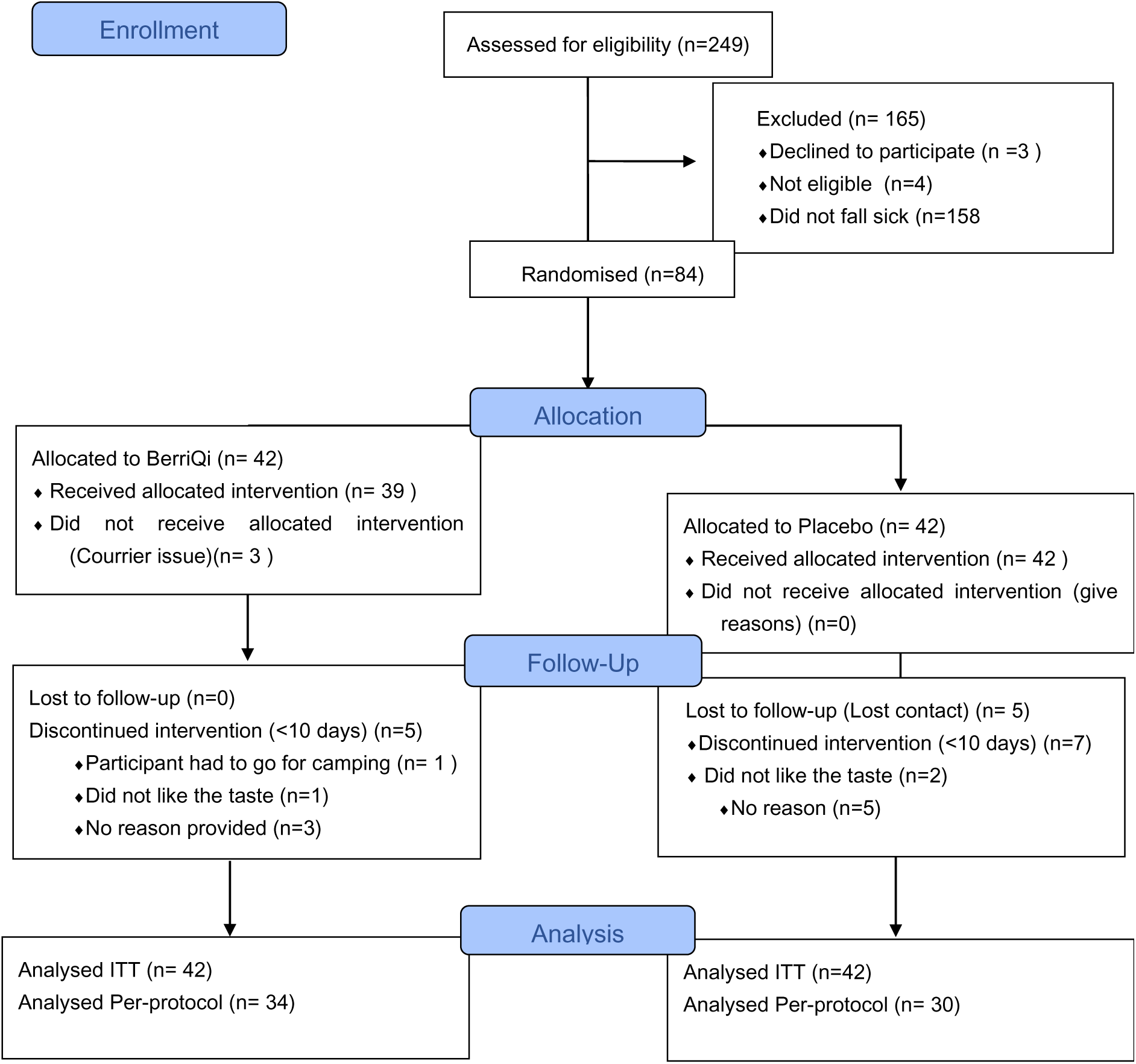
Consort flow diagram describing the recruitment process

Table 1 shows that randomisation produced comparable groups at baseline. This table includes all randomised participants (ITT), regardless of whether they received product or completed diaries.

**Table 1.** Baseline characteristics of study participants.

| Baseline characteristic | Overall<br>N = 84<sup>1</sup> | BerriQi<br>N = 42<sup>1</sup> | Placebo<br>N = 42<sup>1</sup> | P-value<sup>2</sup> |
| --- | --- | --- | --- | --- |
| Age (years) | 8.7 ± 2.9 | 8.7 ± 2.7 | 8.7 ± 3.1 | >0.9 |
| sex |  |  |  | >0.9 |
| Male | 38 (45%) | 19 (45%) | 19 (45%) |  |
| Female | 46 (55%) | 23 (55%) | 23 (55%) |  |
| ethnicity |  |  |  | 0.038 |
| NZ European | 48 (59%) | 23 (58%) | 25 (60%) |  |
| Māori | 19 (23%) | 10 (25%) | 9 (21%) |  |
| Pacific | 4 (4.9%) | 0 (0%) | 4 (9.5%) |  |
| Asian | 6 (7.3%) | 2 (5.0%) | 4 (9.5%) |  |
| Other | 5 (6.1%) | 5 (13%) | 0 (0%) |  |
| Unknown | 2 | 2 | 0 |  |
| Allergy |  |  |  | >0.9 |
| Yes | 3 (3.6%) | 2 (4.8%) | 1 (2.4%) |  |
| No | 81 (96%) | 40 (95%) | 41 (98%) |  |
| Chronic_respiratory issue |  |  |  | 0.4 |
| Yes | 21 (25%) | 9 (21%) | 12 (29%) |  |
| No | 63 (75%) | 33 (79%) | 30 (71%) |  |

### 3.2 Baseline demographic information

Baseline sociodemographic characteristics, including ethnicity, gender, age, and pre-existing respiratory conditions, are included in Table 1. Analyses confirmed that the groups were similar, with no statistically significant differences between the groups in age and gender. The major ethnic groups in the study were New Zealand European and Maori, which were similarly distributed between the two groups. The significant difference in ethnicity between the groups (P=0.038) was due to the distribution of participants in the “Other” ethnicity category (Table 1).

### 3.3 Outcomes measures

#### 3.3.1 Daily global illness scores

The mean global illness severity score decreased steadily in both groups over the 14- day follow-up (Figure 2 A). In the intention-to-treat (ITT) population, there was no significant treatment-by-day interaction (P=0.99) or overall treatment effect (P=0.21). However, the BerriQi group consistently reported lower symptom severity across all days, with a lower overall mean score compared with placebo (1.6 ± 0.7 vs 1.8 ± 0.8) Figure 2 A and D. In the per-protocol (PP) population, restricted to participants with ≥ 80% compliance, a trend toward a treatment effect was observed (P=0.07; Supplementary Figure 2). Analysis of daily scores showed a significant reduction in symptoms in the BerriQi group compared with placebo by day 5 (P =0.04). However, after adjustment for multiple comparisons using the false discovery rate (FDR) across all days, this difference was no longer statistically significant (P=0.580, Supplementary Table 1.

**Figure 2.**
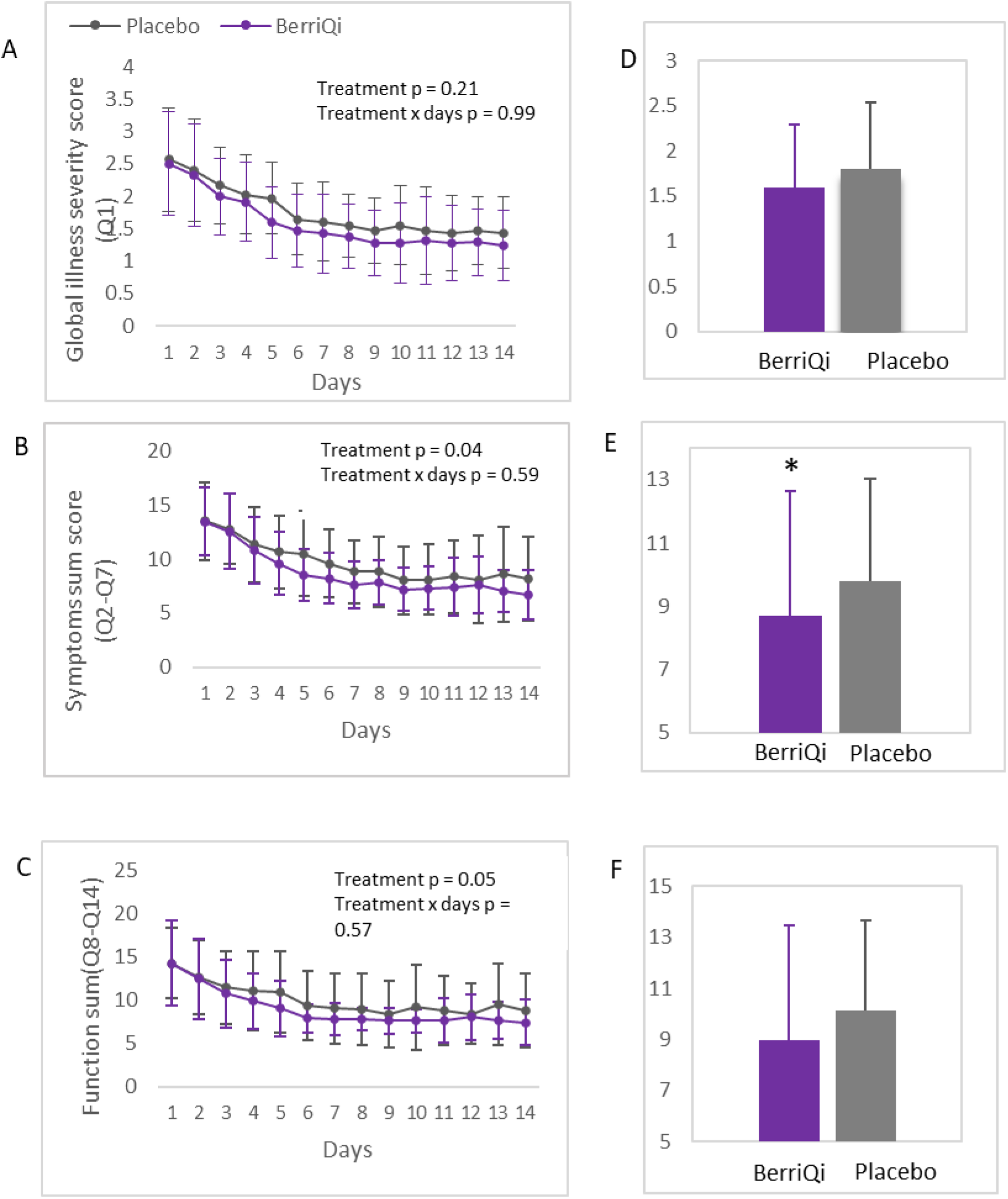
Mean illness severity score over 14 days after intervention (ITT), n = 84. A. Global illness severity score (Q1), B. Symptoms sum score (Q2-Q7) and C. Function sum score (Q8-Q14). Data are presented as means ± SD. D, E, F indicates total treatment effect for global illness severity score, symptoms sum score and function sum respectively.

#### 3.3.2 Symptoms sum (Q2-Q7)

The mean symptom score (runny nose, nasal congestion, sneezing, sore throat, cough, and fatigue) decreased progressively over the 14 days in both groups. In the mixed-effects model, the symptom sum score showed a significant treatment effect (P=0.04) with lower overall symptom severity observed in the BerriQi group compared with placebo (8.7 ± 3.2 vs 9.8 ± 3.9) in the ITT population (Figure 2 B and E), but no significant treatment-by-day interaction was detected (P=0.59). PP subgroup analyses showed comparable findings (Supplementary Figure 2). When symptom scores were analysed on a daily basis, a significant reduction was observed on day 5 in the BerriQi group compared with placebo (P=0.005) which was consistent with the global illness severity score (Supplementary Table 1). However, after adjustment for multiple comparisons using FDR, this result showed only a trend toward significance (P=0.076; Supplementary Table 1). Considering individual symptoms- sneezing, sore throat, stuffy nose and feeling tired showed significantly lower scores in the BerriQi group compared with placebo (Supplementary Figure 3). After adjustment for multiple comparisons using FDR, this result was not significant.

#### 3.3.3 Functions (Q8-Q14)

The mean function score (thinking, sleeping, breathing, talking, walking, going to school, and playing) decreased progressively over the 14 days in both groups. The mixed-effects model indicated lower scores in the BerriQi group, with a trend towards a treatment effect (P=0.05) but no significant treatment-by-day interaction (P=0.57) (Figure 2 C). In the PP subgroup, a significant treatment effect was observed (P=0.02) (Supplementary Figure 2).

#### 3.3.4 Total sick days

Total sick days were calculated using two pre-specified definitions of symptom resolution: (1) a global illness score = 1 (“not sick”), and (2) a symptom sum = 6 (all measured symptoms rated as "not sick). Total sick days were analysed using negative binomial regression.

### Global score =1

Using the global score definition, the per-protocol (PP) analysis showed that the BerriQi group experienced one fewer sick day (median 6 days; 95% CI: 3-9) compared with placebo (median 7 days; 95% CI: 6-11). The incidence rate ratio (IRR) was 0.78 (95% CI: 0.60–1.01), indicating a 22% reduction in sick days; however, this did not reach statistical significance but represented a trend (P = 0.06). The ITT analysis showed similar results, but the P-value was not significant (IRR=0.87 ,95% CI: 0.65- 1.16; P = 0.35).

### Symptoms Sum= 6

Using the symptom sum = 6 definition, the PP analysis demonstrated a significant reduction (P =0.01) in total sick days in the BerriQi group (median 8 days; 95% CI: 4 -12) compared with placebo (median 11 days; 95% CI: 8-13; IRR = 0.79. The ITT group also showed a 3-day reduction but did not reach statistical significance (IRR=0.89 ,95% CI: 0.71-1.12; P=0.32).

### Time to resolution

Time to resolution was analysed by Kaplan–Meier curves, the log-rank test, and a Cox proportional-hazards model, reported as hazard ratios (HR [95% CI]). If participants reported no symptoms for two consecutive days, symptoms were considered resolved.

Symptoms resolved faster in the BerriQi group indicating a clear positive trend towards BerriQi compared with placebo across all the symptoms measured. Functional symptom resolution reached statistical significance in the ITT group: BerriQi-treated children were 79% more likely to achieve functional recovery at any given time point (HR=1.79, 95% CI: 1.09–2.91, P=0.02). (Table 2)

**Table 2.** Time to resolve symptoms.

| Outcome | Time to resolve (Days) |  | Log rank (p value) | Cox HR (95% CI) |
| --- | --- | --- | --- | --- |
|  | BerriQi (Median) | Placebo (Median) |  |  |
| Global illness severity | 5.5 | 6 | 0.08 | 1.54 (0.93–2.56) |
| Symptoms sum (Q2–Q7) | 6 | 9.5 | 0.17 | 1.49 (0.84–2.66) |
| Function sum (Q8–Q14) | 6 | 7 | 0.02 | 1.79 (1.09–2.91) |

Time to resolve the symptoms is based on reporting of no symptoms for 2 consecutive days. Data are presented for Intention to Treat (ITT) n = 84

## 4 Discussion

This randomised, double-blind, placebo-controlled trial provides the first clinical evidence that a whole-fruit Boysenberry and apple blend (BerriQi) significantly alleviates URTI symptoms in children. The global illness score declined steadily over the study period, reflecting progressive symptom improvement, however, this did not reach statistical significance. Nevetheless, BerriQi significantly reduced the number of sick days, shortening the duration and severity of common cold symptoms, thereby supporting children’s return to normal daily function. As URTI remains the leading cause of school absenteeism globally, these findings suggest that daily BerriQi intake may help reduce this burden.

BerriQi has previously demonstrated *in vivo* efficacy in reducing mucus production and airway tissue damage due to inflammation, which results in improved lung capacity and function (Shaw et al., 2021). The resolution of inflammation is mediated by alveolar macrophages, which change their action depending on the pathogen and phase of infection (Byrne et al., 2015). During respiratory infection, macrophages adopt a pro-inflammatory state (M1) in which they recruit additional immune cells, produce reactive oxygen species, and release pro-inflammatory cytokines to fight the pathogen (Forman & Torres, 2002; Martinez et al., 2008). Once the pathogen is cleared, recovery relies on macrophages switching to a tissue-repair state (M2), producing anti-inflammatory cytokines and depositing collagen at the sites of mucosal damage. After tissue repair, macrophages enter a clean-up state (M2C), which breaks downcollagen, allowing normal airway function to be restored (Russell et al., 2002; Lim et al., 2000). When this macrophage switching sequence is delayed or incomplete, the pro-inflammatory M1 and collagen-intensive M2 states persist longer than necessary, maintaining the conditions that produce ongoing mucus, congestion, and fatigue even after the infection itself has resolved, resulting in lingering symptoms after infection. BerriQi has been shown to actively trigger this macrophage switching from M1 into the M2 and further into the M2C recovery states, with corresponding reductions in immune cell infiltration, excess collagen, and mucus production in airway tissue (Shaw et al., 2016, 2021).This activity is attributed to the unique polyphenol profile of BerriQi, particularly the complementary activities of procyanidins, anthocyanins and quercetin. Anthocyanins mainly come from the boysenberries, but quercetin and procyanidins are derived from both apples and boysenberries. The unique ratio of Boysenberries and apples in BerriQi provides a synergistic effect which is not demonstrated by Boysenberries or apples alone (Shaw & Hurst, 2019).

The preclinical rationale described by Shaw & Hurst directly guided the placebo design of the present study. Rather than an inert placebo, children in the placebo group received a heat-treated Boysenberry and apple — a matrix that preserves the full complement of heat-stable nutrients, vitamins, and minerals naturally present in Boysenberry while selectively degrading the heat-sensitive fraction of the polyphenols. The total dose of actives (anthocyanins, quercetin, and procyanidins) delivered in the BerriQi formulation was 6.4 mg per day (two chewable tablets), compared with 0.5 mg per day (two chewable tablets) in the placebo. This design was chosen deliberately; given that participants were acutely unwell children, it was considered ethically appropriate to provide a placebo that retained the general nutritional benefits of whole fruit rather than an entirely inert placebo. Had a fully inert placebo been used, the magnitude of the observed treatment effect would likely have been greater. The clinical outcomes reported here therefore represent a conservative estimate of BerriQi’s true benefit in paediatric URTI.

While the intention-to-treat (ITT) analysis served as the primary outcome measure, the per-protocol (PP) analysis, restricted to compliant participants, yielded more pronounced treatment effects, highlighting the importance of consistent BerriQi consumption in achieving observable clinical benefits. Notably, the overall pattern of results remained consistent across both analytical approaches. All parameters, including mean global illness severity scores, symptom scores, and function scores, declined steadily over time in each group. However, a statistically significant between-group difference emerged at day 5, with the BerriQi group demonstrating markedly lower values compared to placebo. This finding suggests that early intervention may be critical in attenuating symptom burden and potentially reducing total sick days. Nevertheless, these results should be interpreted with caution, as false discovery rate (FDR)-adjusted comparisons did not reach statistical significance. Therefore, further studies are warranted in larger cohorts to substantiate these findings.

The benefits of BerriQi in reducing respiratory symptoms have been previously demonstrated following exposures to environmental stressors such as ozone (Lomiwes et al., 2025). While Lomiwes et al reported that a BerriQi intervention for 5 days improved respiratory symptoms such as throat irritation after ozone exposure, this is the first study to explore its effect on URTI symptoms in children. This study provides evidence for the benefits of BerriQi in reducing URTI-related symptoms in children, but a few limitations warrant consideration. The study design presented logistical challenges, particularly regarding timely investigational product (IP) delivery, as participants were recruited from across New Zealand and were required to receive the IP within 24–48 hours of symptom onset. Consequently, three participants did not receive the product within the stipulated timeframe. Quantification of school absenteeism was complicated by scheduled holidays and weekends, and sick days were therefore estimated using negative binomial regression based on two criteria: the global illness score, reflecting overall wellbeing, and the composite symptom score, capturing UTRI-specific symptoms. Symptom resolution was conservatively defined as two consecutive symptom-free days, minimising misclassification and enhancing analytical rigour. Furthermore, compliance was suboptimal in a subset of participants due to poor acceptability of the chewable formulation’s taste and texture, leading to reduced adherence or voluntary withdrawal. Additionally, inconsistent caregiver reporting resulted in missing symptom data. Despite these limitations, overall compliance exceeded 80%, providing sufficient statistical power and supporting the robustness of the study findings. Nevertheless, a larger sample size, as well as studies in adults, would have further strengthened the interpretability and generalisability of the findings. Future studies in larger cohorts and across a broader range of age groups are therefore warranted.

## 5 Conclusions

The findings from the present study suggest that consistent intake of BerriQi may contribute to faster and more complete recovery from mild respiratory illness. BerriQi may be considered a safe and natural paediatric supplement that may help alleviate symptoms and support faster recovery from respiratory illness.

## Data Availability

The datasets generated during and/or analysed during the current study are available from the corresponding author on reasonable request

## 6 Declarations

### Trial registration

The trial was registered with the Australia New Zealand Clinical Trials Registry (registration number ACTRN: 12624000398505) (http://www.anzctr.org.au/).

Competing interests: AS, SM, EG were employees of Anagenix IP Ltd that manufactures and sells BerriQi powder, at the time of the study.

### Funding

This study received funding from Anagenix Ltd

### Ethics approval and consent to participate

The study was conducted in accordance with the Declaration of Helsinki and approved by New Zealand’s Southern Health and Disability Ethics Committee (18th July 2024).

### Author Contributions

AS: designed study, ethics and protocol development, conducted the study, wrote-original draft, reviewed and edited manuscript and had primary responsibility for the final content; EG: Designed the study, conducted research, reviewed, and edited manuscript; and SM designed the study, ethics and protocol development, reviewed and edited manuscript. All authors approve the final version of the manuscript for submission.

## Acknowledgments

The authors gratefully acknowledge University of Auckland for providing data collection and analysis support. Data collection and analysis was done independently by Data management and analysis team from University of Auckland. Logistics team for blinding and delivery of IP. Also, the authors would like to acknowledge participants and the caregivers for their time and participation during the trial.

global illness severity score, symptoms sum score and function sum respectively.

## Notes

### Clinical Trial

ACTRN: 12624000398505

### Author Declarations

The study was approved by New Zealands Southern Health and Disability Ethics Committee

