## Supplemental Table 1 for "Effect of Boysenberry apple powder blend (BerriQi^®^) on reducing symptom severity in children with Upper respiratory tract infection"

**Supplementary Table 1**. Day to day reporting of symptoms WURSS-k Survey

| | | ITT | | PP | |
|---|---|---|---|---|---|
| **Outcome** | **Day** | P value | P-value (FDR across days) | P value | P-value (FDR across days) |
| Global illness severity (Q1) | 1 | 0.704 | 0.758 | 0.455 | 0.530 |
| | 2 | 0.950 | 0.950 | 0.696 | 0.696 |
| | 3 | 0.420 | 0.758 | 0.376 | 0.479 |
| | 4 | 0.662 | 0.758 | 0.292 | 0.479 |
| | 5 | 0.042 | 0.589 | 0.012 | 0.167 |
| | 6 | 0.340 | 0.758 | 0.208 | 0.479 |
| | 7 | 0.414 | 0.758 | 0.259 | 0.479 |
| | 8 | 0.558 | 0.758 | 0.353 | 0.479 |
| | 9 | 0.496 | 0.758 | 0.363 | 0.479 |
| | 10 | 0.225 | 0.758 | 0.117 | 0.479 |
| | 11 | 0.693 | 0.758 | 0.520 | 0.561 |
| | 12 | 0.445 | 0.758 | 0.276 | 0.479 |
| | 13 | 0.413 | 0.758 | 0.235 | 0.479 |
| | 14 | 0.462 | 0.758 | 0.180 | 0.479 |
| Symptoms (Q2-Q7) | 1 | 0.616 | 0.670 | 0.346 | 0.437 |
| | 2 | 0.698 | 0.698 | 0.774 | 0.774 |
| | 3 | 0.425 | 0.541 | 0.375 | 0.437 |
| | 4 | 0.156 | 0.363 | 0.143 | 0.324 |
| | 5 | 0.005 | 0.076 | 0.004 | 0.061 |
| | 6 | 0.050 | 0.175 | 0.041 | 0.157 |
| | 7 | 0.134 | 0.363 | 0.125 | 0.324 |
| | 8 | 0.227 | 0.437 | 0.189 | 0.324 |
| | 9 | 0.281 | 0.437 | 0.195 | 0.324 |
| | 10 | 0.325 | 0.455 | 0.300 | 0.420 |
| | 11 | 0.276 | 0.437 | 0.208 | 0.324 |
| | 12 | 0.622 | 0.670 | 0.626 | 0.674 |
| | 13 | 0.046 | 0.175 | 0.045 | 0.157 |
| | 14 | 0.032 | 0.175 | 0.022 | 0.155 |
| Functions (Q8-Q14) | 1 | 0.868 | 0.935 | 0.790 | 0.851 |
| | 2 | 0.967 | 0.967 | 0.878 | 0.878 |
| | 3 | 0.530 | 0.674 | 0.666 | 0.847 |
| | 4 | 0.182 | 0.320 | 0.161 | 0.282 |
| | 5 | 0.034 | 0.240 | 0.058 | 0.263 |
| | 6 | 0.069 | 0.275 | 0.081 | 0.263 |
| | 7 | 0.152 | 0.320 | 0.149 | 0.282 |
| | 8 | 0.183 | 0.320 | 0.159 | 0.282 |
| | 9 | 0.394 | 0.552 | 0.286 | 0.401 |
| | 10 | 0.079 | 0.275 | 0.094 | 0.263 |
| | 11 | 0.271 | 0.421 | 0.243 | 0.378 |
| | 12 | 0.731 | 0.852 | 0.769 | 0.851 |
| | 13 | 0.033 | 0.240 | 0.062 | 0.263 |
| | 14 | 0.103 | 0.289 | 0.090 | 0.263 |
