## Supplementary Figure 1 for "Effect of Boysenberry apple powder blend (BerriQi^®^) on reducing symptom severity in children with Upper respiratory tract infection"

### Wisconsin Upper Respiratory Symptom Survey for kids—daily symptom report

| **Day:** | **Date:** | **Time:** | **ID:** |
|---|---|---|---|

**Please fill in one circle for each question:**

| | Not sick | A little sick | Sick | Very sick |
|---|---|---|---|---|
| How sick do you feel today? | O | O | O | O |

**How bad are your cold symptoms?** (Overall, since yesterday)

| | Do not have this | A little bad | Bad | Very bad |
|---|---|---|---|---|
| Runny nose | O | O | O | O |
| Stuffy nose | O | O | O | O |
| Sneezing | O | O | O | O |
| Sore throat (hurts to swallow) | O | O | O | O |
| Cough | O | O | O | O |
| Feeling tired | O | O | O | O |

**Since yesterday**, how **hard** has it been to:

| | Not at all | A little hard | Hard | Very hard |
|---|---|---|---|---|
| Think | O | O | O | O |
| Sleep | O | O | O | O |
| Breathe | O | O | O | O |
| Talk | O | O | O | O |
| Walk, climb stairs, exercise | O | O | O | O |
| Go to school | O | O | O | O |
| Play with friends | O | O | O | O |

**Compared to yesterday**, I feel my cold is...

| A lot better | A little better | The same | A little worse | A lot worse |
|---|---|---|---|---|
| O | O | O | O | O |

**I completed this page:** ☐ all by myself ☐ with some help ☐ with a lot of help

Who helped you? ____________________________
