## Supplementary Figure 2 for "Effect of Boysenberry apple powder blend (BerriQi^®^) on reducing symptom severity in children with Upper respiratory tract infection"

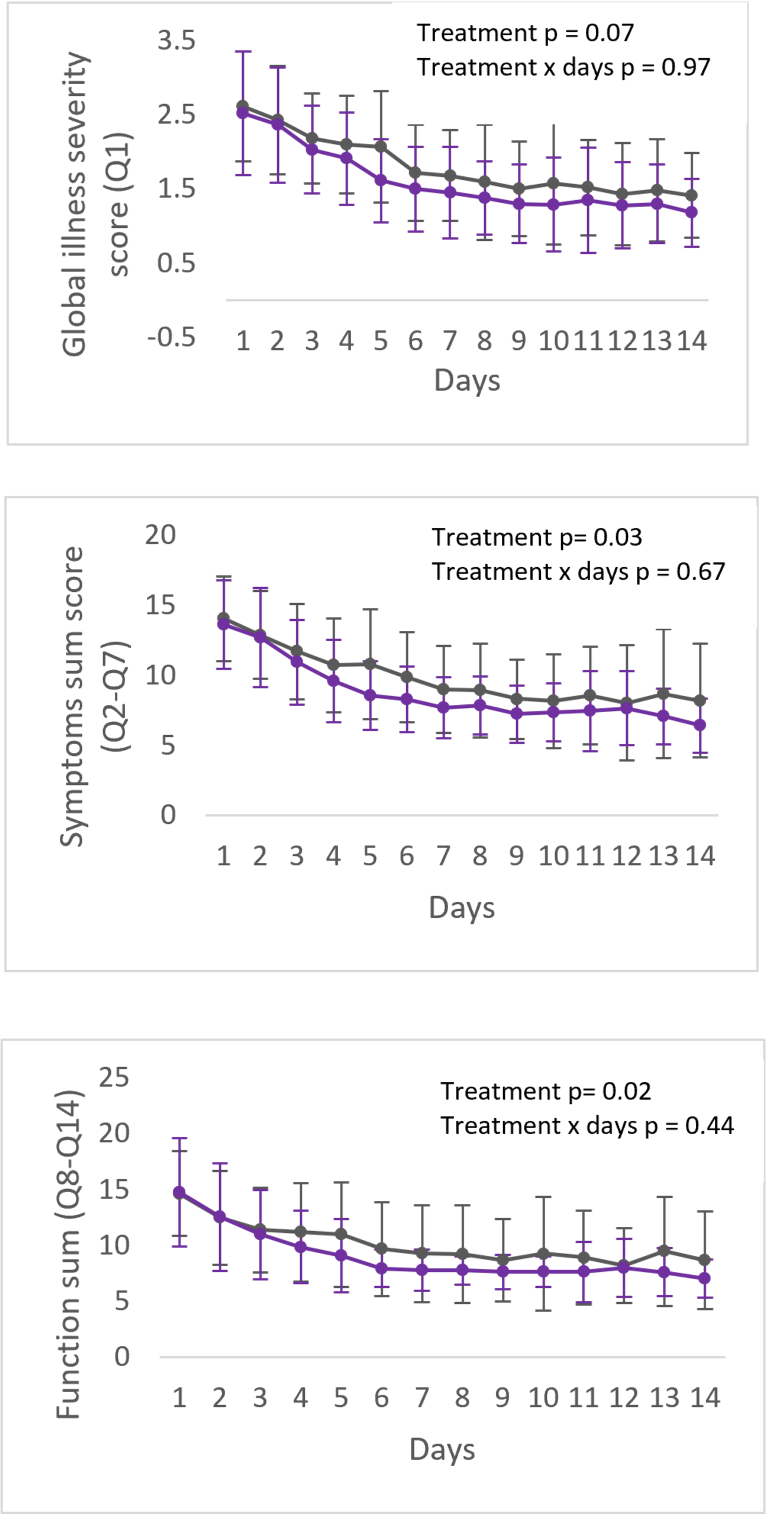


*Supplementary Figure 2: Mean illness severity score over 14 days after intervention (PP), n = 64. A. Global illness severity score (Q1), B. Symptoms sum score (Q2-Q7) and C. Function sum score (Q8-Q14). Data are presented as means ± SD.*
