## Supplementary figures and images for "Effect of Boysenberry apple powder blend (BerriQi^®^) on reducing symptom severity in children with Upper respiratory tract infection"

### Supplementary Figure 3

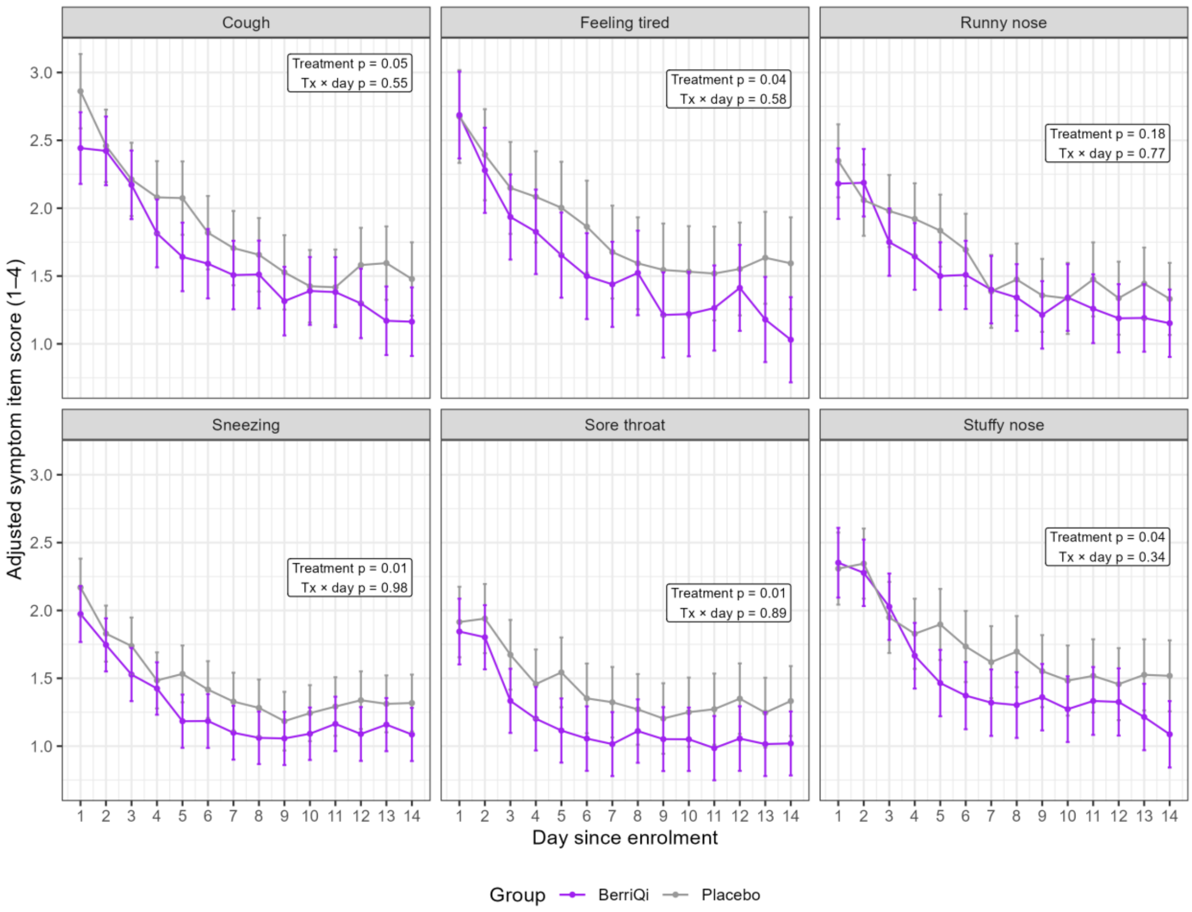


*Supplementary figure 3: Individual symptoms severity (Q2-Q7) over 14 days after intervention (ITT), N=84*
